# The Effect of Short Message Service (SMS)-Reminders on Child Health in Parents of Newborns: A Pilot Randomised Controlled Trial

**DOI:** 10.1101/2021.01.12.21249639

**Authors:** M Shah, J Ramsay, M Dymock, J Marsh, N Newall, C McCallum, J Davis, T Snelling

## Abstract

**Background:** Incomplete vaccination in Australia is greatly due to vaccine hesitancy, which is driven by multiple factors. Studies analysing the effect of behavioural ‘nudges’ on vaccine uptake have shown promising results. Although there is some evidence for positively framed SMS vaccine messages, evidence is lacking for loss-based framed messages which may be highly effective but may paradoxically risk increasing vaccine hesitancy. This pilot study aimed to evaluate the effect of loss-based framed SMS messages on vaccine hesitancy.

**Methods:** This single-blinded randomised controlled trial enrolled pregnant women from antenatal clinics in King Edward Memorial Hospital, Perth, WA which delivers approximately 6000 higher-risk infants per year. Participants were randomised to receive either a loss-based framed SMS message warning of the risks of failure to vaccinate, or a sham SMS message unrelated to vaccination, and were asked to complete the short-scale vaccine hesitancy questionnaire before and after the intervention.

**Discussion:** The application of behavioural sciences through SMS-reminders has the potential to improve vaccination rates. There are difficulties in engaging with parents of newborns about vaccine hesitancy. Further research is warranted using other approaches to recruitment.

**Highlights:**

- Pilot randomised controlled trial on the use of SMS reminders to influence vaccine hesitancy
- Application of behavioural economics, nudge techniques and mobile health
- Vaccine hesitancy measured using the short-scale PACV questionnaire
- There are difficulties in engaging with parents of newborns about vaccine hesitancy
- Further research needed to optimise message framing, timing and reduce vaccine hesitancy

**Trial Registration:** The trial is registered with the Australia and New Zealand Clinical Trials Register (ACTRN12618001510235).

## Introduction

Vaccinating children greatly reduces the risk of a number of targeted infectious diseases. [1] Vaccinated populations also benefit from herd immunity, where the risk of an individual contracting an infection is reduced because most or all of their contacts are immune. [2] It’s generally thought that up to 95% of the population need to be vaccinated to achieve effective herd immunity. [3, 4] In Australia, no jurisdiction consistently achieves vaccine coverage above this rate, [5] even for early childhood vaccines, leading to an avoidable risk of morbidity and mortality from these preventable diseases. [3]

‘Vaccine hesitancy’ is defined as a delay in acceptance of vaccines despite their availability. [6, 7] This occurs on a spectrum, and is influenced by confidence in the vaccine provider, the perceived need of the vaccine, its potential side effects, and complexity of vaccination schedules. [4, 8, 9] Strategies targeting vaccine hesitancy among parents of newborns might improve vaccination rates and health outcomes. Effective strategies may be informed by the field of behavioural economics, a field of research that combines behavioural science with economic reasoning. [10] The objective of such strategies is to encourage or ‘nudge’ individuals toward a positive behaviour, without resorting to coercion.[11] Bickel *et al*. argue this approach should be based on a scientific understanding of health behaviours (to inform practical interventions) and should make use of available technology to maximise efficacy and cost-effectiveness. [12] This non-coercive approach might prove more effective and acceptable than regulation or punitive strategies. [7, 13, 14]

Behavioural nudges can be implemented as SMS messages. With more than 80% of adult Australians owning a mobile phone, delivering nudges by SMS may be simple and efficient for personally tailoring preventative health behaviour modification. [15-18] SMS nudges have shown promise in studies aiming to promote lifestyle changes in people with chronic diseases and have been shown to be more effective than phone calls for increasing vaccine uptake among parents and adolescents. [19, 20] A recent study also showed positive effects of smart phone based interventions on the health behaviours of new mothers, directly translated to better infant health. [21]

The use of SMS to improve childhood vaccination rates is an active area of research. A systematic review of the use of SMS-messages as preventative health interventions noted that the framing of the messages may be important. [15] Positive, motivational and ‘gains-based’ framing of messages emphasise the benefits of a specific behaviour; they have been found to be effective for encouraging preventative health in some contexts. [22-24] Negative or ‘loss-based’ framing emphasise instead the risks associated with failure to adopt the desired behaviour; such messaging may be more effective for deterring high risk behaviours. [24-26] A meta-analysis comparing the effectiveness of gains vs loss-based framing showed no clear advantage of using either technique for promoting preventative health behaviours.[26] The effects of loss-based framing may be temporary and may even paradoxically deter positive health behaviours in some contexts.[25, 27] In a recent experiment, presenting vaccine hesitant parents with images or narratives of children affected by measles appeared to make them more concerned about vaccine side effects. We are planning a large multi-arm trial of SMS reminders for improving the timeliness of routine vaccination of young children(ACTRN12618000789268). Given the potential for double-edged effects of loss-based SMS reminders, we aimed to first pilot their use in a small group of new parents by assessing for any effect on vaccine hesitancy compared to a sham SMS-message unrelated to vaccination.

## Materials and Methods

### Design

A pilot single-blind randomised controlled trial of SMS messages targeting vaccine hesitancy in new parents. A timeline of study procedures is described by **Figure 1**.

**Figure 1:**
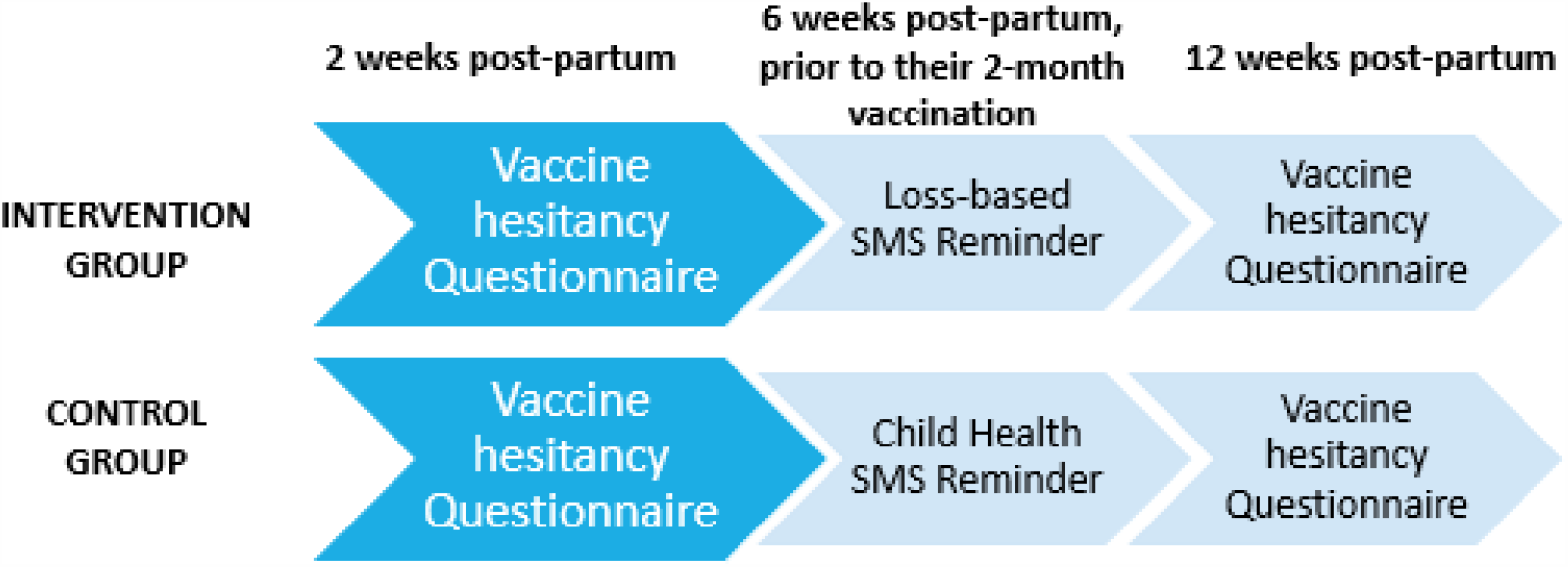
SMS-Reminder Design

### Study Population and Recruitment

We enrolled parents presenting for antenatal care at a large tertiary maternity hospital in Perth, Western Australia which attends to more than 6000 deliveries per year.[28] Expecting parents meeting the eligibility criteria (described below) were told that the purpose of the study was to evaluate the use of parental SMS messages in child health; they were invited to consent for follow-up contact in the postpartum period. We did not explicitly reveal our objective of assessing the impact of loss-based framed SMS messages on vaccine hesitancy to minimise the risk of selection bias.

### Participant eligibility

We included parents expecting their first child, 18 years or older, English-speaking, ≥28 weeks pregnant at first contact, owning a mobile phone with text messaging and data/Wi-Fi connection capability, and willing to be contacted via mobile phone. We excluded parents unable to give informed consent and/or comply with the study or otherwise deemed unsuitable by the clinic staff. Parents of stillborn infants and those delivering before 38 completed weeks of gestation were eliminated prior to randomisation.

### Intervention and Follow-up

Approximately two weeks after delivery, parents were recontacted via an SMS-message containing a link to an online consent form to confirm their participation. They were then directed to complete a vaccine hesitancy survey.

Participants responding to the survey were then randomised to receive either a loss framed SMS message to encourage vaccination, or a sham health-related SMS message (unrelated to vaccination) at approximately six weeks after delivery, just before the first vaccine dose is scheduled to be given. In Australia the 2-month schedule point comprises a 6-in-1 DTPa, hepatitis B, polio, *Haemophilus influenzae* type B vaccine, a multivalent conjugated pneumococcal vaccine, and an oral rotavirus vaccine. (28). All SMS messages were sent during daytime hours.

The SMS message sent to the intervention group read: ‘Telethon Kids health message: Your child is now due for their 2-month vaccination. Delaying a child’s vaccination can put them and other children at risk. Do not reply to this text message.’ This message framing was informed by a community reference group of parents with young children; the group’s preferred loss-based message was selected for this study.

The sham SMS health message sent to the control group read: ‘Telethon Kids health message: Sleeping your baby on his or her back is safest for preventing SIDS (Sudden Infant Death Syndrome). Do not reply to this text message.’

Participants were then sent a follow-up vaccine-hesitancy survey approximately 12 weeks after delivery. Reminders for completion of both surveys were sent at weekly intervals for up to 3 weeks, after which an attempt was made to contact non-responders by telephone to confirm their continued participation and, if applicable, to verbally complete the questionnaire.

### Vaccine hesitancy survey

Surveys were sent electronically via SMS message and self-administered. Responses were categorical, and the survey tool included in-built logic checks to ensure complete data entry. A shortened form of the Parental Attitudes towards Childhood Vaccination (PACV) questionnaire was used.[29] The original PACV has been validated to ensure content validity[29]; the short form has been validated and found to be similar to the original for identifying and classifying parental hesitancy.[30] We included five additional questions related to general child health to mask the objective of the study. Some terminology was modified to better align with Australian terminology (e.g. ‘shot’ replaced with ‘vaccination’).

The SS-PACV score was calculated from the individual question responses (0, 1 or 2 points each) and summed to a total score from a maximum of 10 points **(Appendix 1)**. This score was compared before and after delivery of the SMS message. Each participant was categorised as having low (score 0-4), medium (score 5-6) or high (score 7-10) vaccine hesitancy at each time point. The primary outcome was the effect of the SMS message as measured by a change in the SS-PACV score and any resultant change in vaccine hesitancy category.

### Statistical analysis

The changes in SS-PACV scores at the post-intervention timepoint (primary outcome) were compared using the Wilcoxon-Mann-Whitney test at a 5% significance level.

### Statistical power

A planned sample size of 274 participants (137 per invention arm) was calculated to enable detection of a 30% lower proportion of participants categorised as low vaccine hesitancy (score 0-4 out of 10) in the intervention compared to the control group. This was based on: (i) expected proportions with low, medium and high vaccine hesitancy in the control arm of 72%, 13% and 15%, respectively[30]; (ii) in the intervention arm, a 30% movement from low to medium vaccine hesitancy category (thus the proportions for the low, medium and high categories would be 50%, 35% and 15%, respectively); (iii) significance level (α) of 5%; (iv) power of 80%; using the sample size formula for the Wilcoxon-Mann-Whitney test by Zhao et al. [31]

## Results

One hundred and eighty-five expecting parents consented to be re-contacted; 45 were subsequently eliminated **(Figure 2)** and 61 agreed to participate and were invited to complete the first PACV survey.

**Figure 2:**
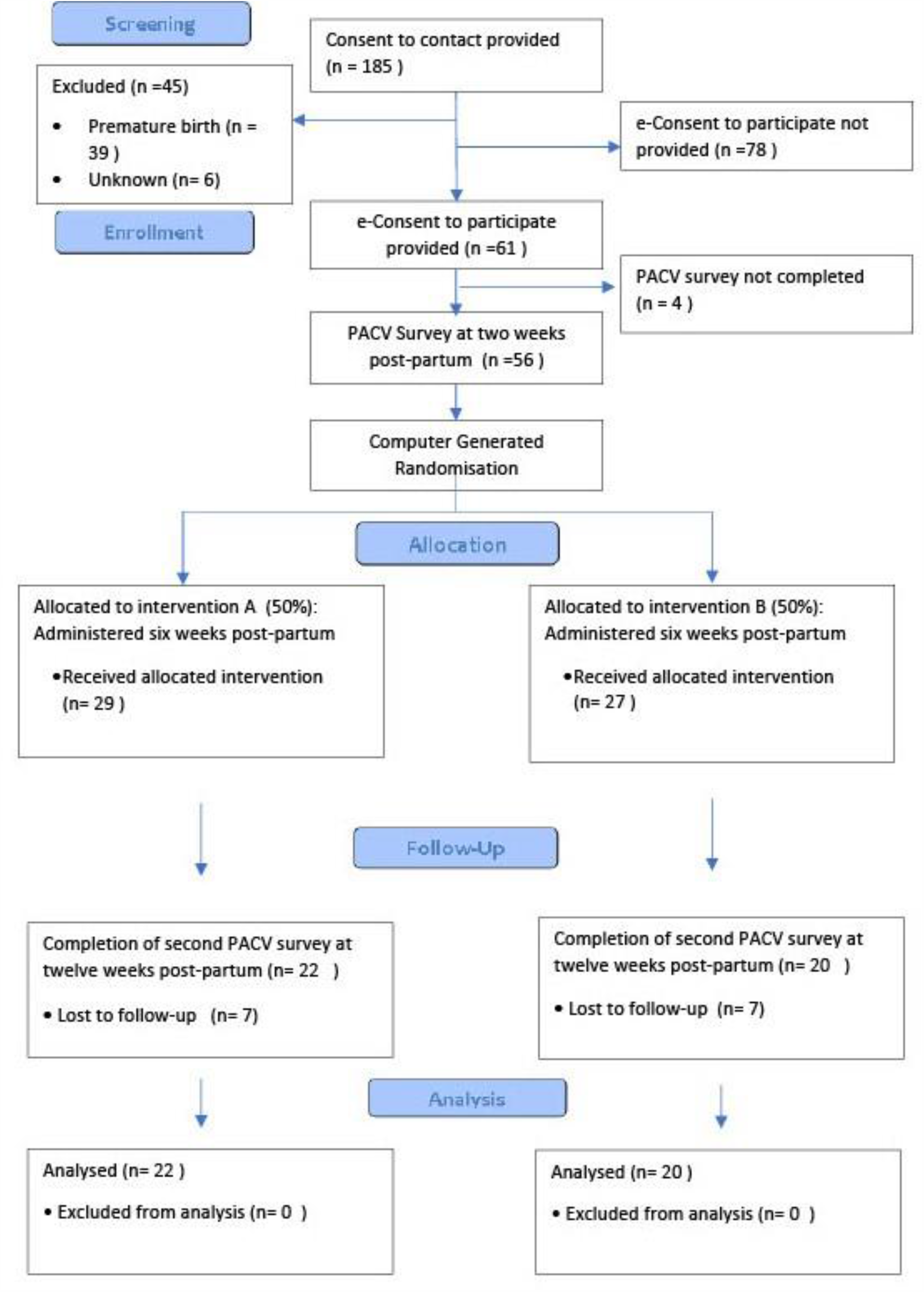
SMS-Reminder Consort Diagram

Of these, 56 completed the first PACV survey and 52 were randomised into the intervention (29) or control group (27). 22 participants in the intervention group and 20 in the control group completed the follow-up survey.

The distributions of the SS-PACV scores before and after the survey and between treatment groups are provided in Figure 3. The median vaccine hesitancy scores changed from 1 (1) at baseline to 1(1.75) at follow-up in the intervention group, and from 1 (1) to 1 (1.25) in the control group. The difference in the change in scores between groups was no greater than expected by chance (p = 0.745). To quantify the probability that the intervention significantly worsened vaccine hesitancy, we conducted a post-hoc Bayesian analysis. We estimated that the probability that the mean change in hesitancy score in the investigation group was more than a unit higher than the mean change in hesitancy score in the control group was 3%.

**Figure 3.**
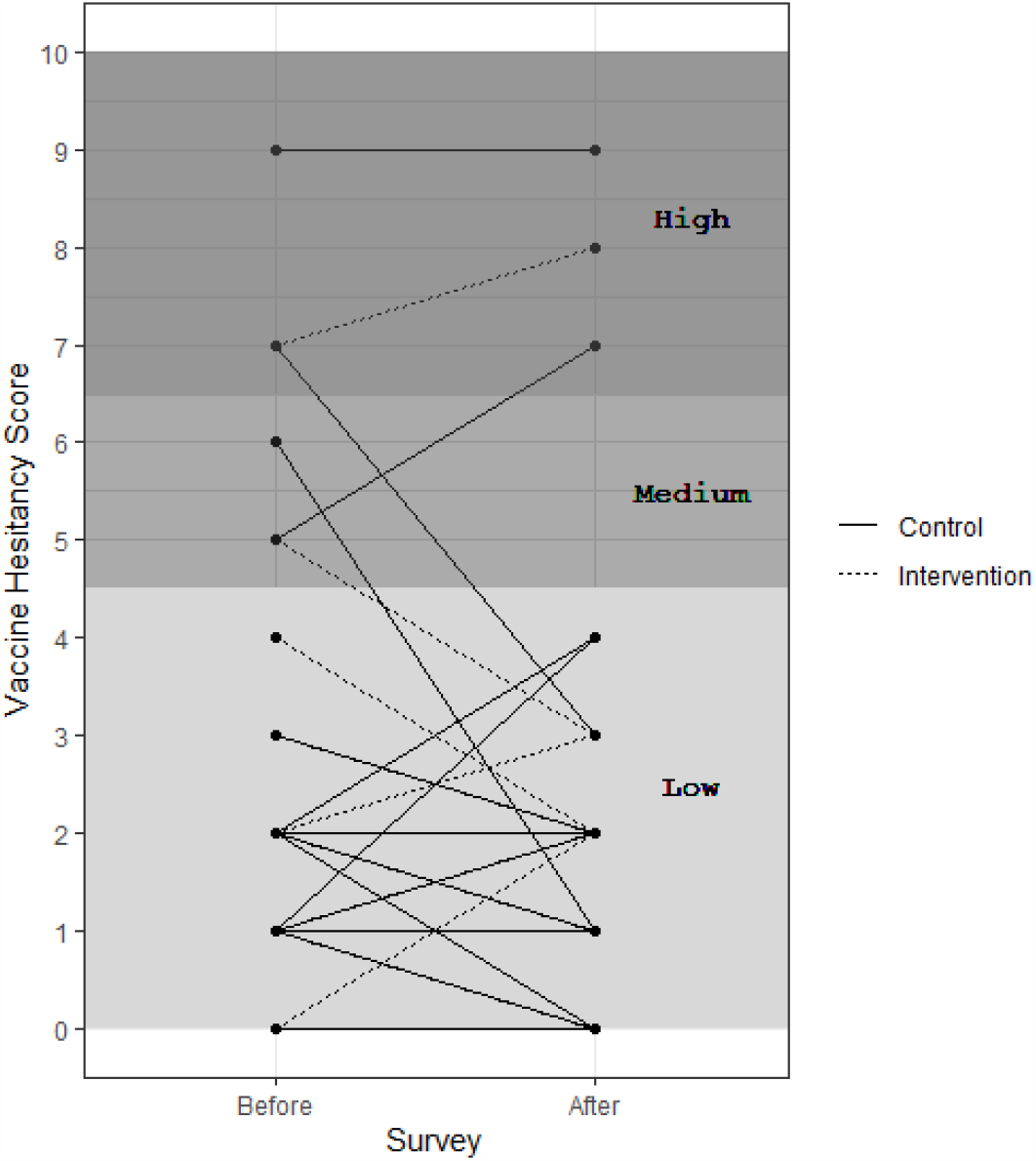
SS-PACV (vaccine hesitancy) scores before and after survey and between treatment allocations. The SS-PACV scores range from 0 (minimum vaccine hesitancy) to 10 (maximum vaccine hesitancy) with categories low (0-4), medium (5-6) and high (7-10).

## Discussion

Vaccine hesitancy is thought to be a major driver of incomplete vaccination in Australia. [4, 5, 7, 8] The use of SMS messaging to deliver ‘nudges’ which target vaccine hesitancy has theoretical promise, but so far there is only limited evidence to support it. [10] Despite concerns about possible paradoxical effects, we found no evidence that loss-based framed SMS-messages increased vaccine hesitancy in parents of newborns, and that an increase in hesitancy, if any, is likely to be small (<1 unit). Through this pilot randomised controlled trial, we were able to identify a number of challenges and design considerations that will need to be addressed before further investigation of the effectiveness of SMS messages for vaccination. Implementation of these changes will enable better powered studies and quality results.

One of the key limitations was slow enrolment of participants despite access to a large potential study population. This was partly due to the eligibility criteria only including parents expecting their first child. The intention of this criterion was to define a group who might be at greatest risk of vaccine hesitancy, and least likely to have attitudes already pre-formed by previous experience (positive or negative) of vaccination and/or infection in previous children. However, this criterion created an impediment to enrolment, resulting in a broadening of the inclusion criteria midway through enrolment. We do not believe this affected the applicability of our findings and future trials should consider the broader inclusion criterion to improve enrolment.

The two-step enrolment procedure, in which parents who were consented in late pregnancy were then re-contacted after delivery, was complex and may have contributed to low participation and completion. Follow-up of these participants was difficult and time-consuming. This may affect the generalisability of the results. Future studies should make participation as easy as possible by requesting ethics approval for a waiver of consent or opt-out consent. We believe that this should not only make enrolment more efficient, it would be ethically justified because the risk of harm is low, and because it would reduce selection bias which favours inclusion of more motivated parents. In addition, future studies should consider measuring vaccination hesitancy and uptake post intervention, to allow for assessment of SMS-reminder efficacy.

## Conclusion

The application of nudge and other techniques from the behavioural sciences are an active area of health research, including for improving vaccination rates. There is a legitimate concern that loss-based framed messages may, paradoxically, induce vaccine hesitancy. Notwithstanding its limitations, our pilot trial found no evidence that loss-based framed SMS-messages are likely to induce vaccine hesitancy. Further research is needed to determine whether such messages are effective, and to identify the optimal message framing and timing for improving vaccine coverage and timeliness and for reducing the morbidity associated with vaccine-preventable diseases. To this end, we have recently commenced AuTOMATIC, a much larger and more pragmatic trial to investigate the effectiveness of loss-based framed messages against usual care and against positively framed and neutral messages, on a whole-of-population basis.

## Data Availability

The datasets used and/or analysed during the current study are available from the corresponding author on reasonable request.

## Abbreviations

SMS: Short-message service
PACV: Parental Attitudes towards Childhood Vaccination
SS-PACV: Short Scale- Parental Attitudes Towards Childhood Vaccination

## Declarations

### Ethics Approval and Consent to Participate

This protocol was approved by the Women and Newborn Health Service Ethics Committee in July 2018. (PRN: RGS0000001045). All aspects of this study were carried out in an ethical manner. Informed consent was obtained after the nature and possible consequences of the study had been fully explained.

### Authors’ Contributions

MS conceptualised the project and drafted the manuscript along with JR, TS and JM. MD analysed and interpreted the participant data. JD, NN and CM provided critical review of the protocol. All authors read and approved the final protocol.

## Acknowledgements

TS holds a Career Development Fellowship from the National Health and Medical Research Council (NHMRC) (GNT 1111657)

---

**Appendix 1:**
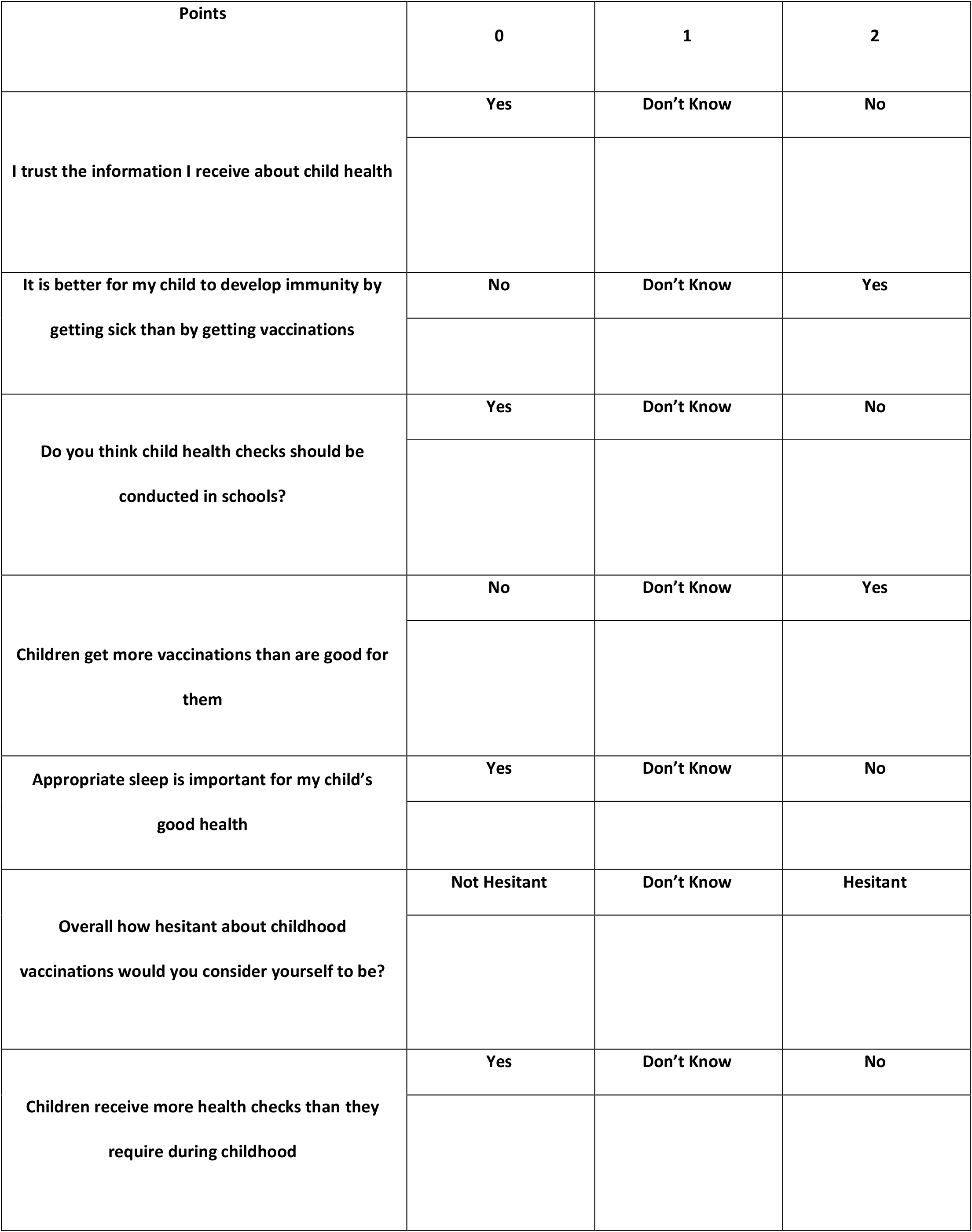
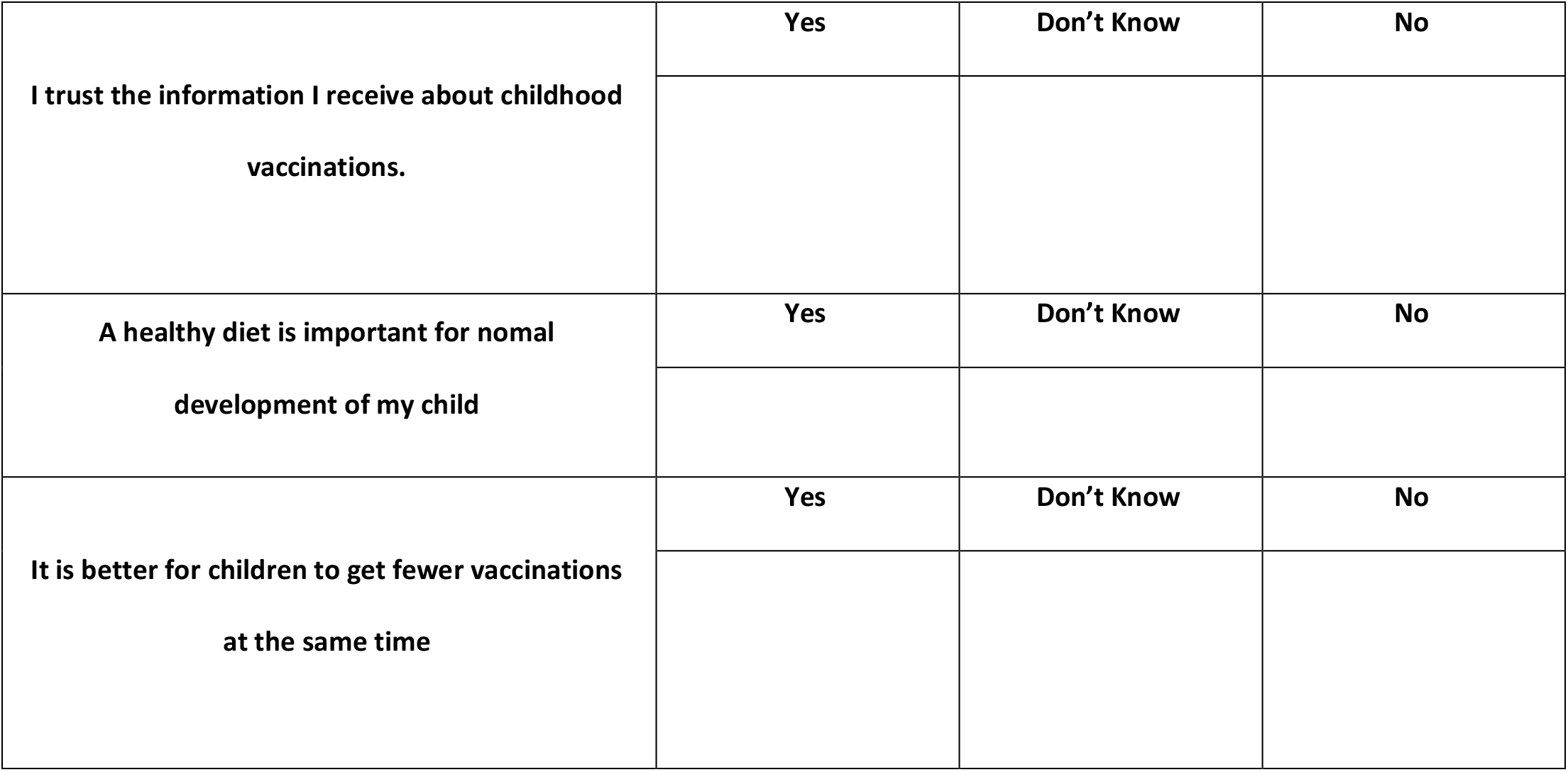
Modified SS-PACV Survey

