## Appendix 1 for "The Effect of Short Message Service (SMS)-Reminders on Child Health in Parents of Newborns: A Pilot Randomised Controlled Trial"

| Points | 0 | 1 | 2 |
|---|---|---|---|
| I trust the information I receive about child health | Yes | Don’t Know | No |
| It is better for my child to develop immunity by getting sick than by getting vaccinations | No | Don’t Know | Yes |
| Do you think child health checks should be conducted in schools? | Yes | Don’t Know | No |
| Children get more vaccinations than are good for them | No | Don’t Know | Yes |
| Appropriate sleep is important for my child’s good health | Yes | Don’t Know | No |
| Overall how hesitant about childhood vaccinations would you consider yourself to be? | Not Hesitant | Don’t Know | Hesitant |
| Children receive more health checks than they require during childhood | Yes | Don’t Know | No |
| I trust the information I receive about childhood vaccinations. | Yes | Don’t Know | No |

| | | | |
|---|---|---|---|
| **A healthy diet is important for nomal development of my child** | **Yes** | **Don't Know** | **No** |
| **It is better for children to get fewer vaccinations at the same time** | **Yes** | **Don't Know** | **No** |
